# Digital Health Misinformation and HPV Vaccine Awareness Among U.S. Adults: A National HINTS Analysis

**DOI:** 10.64898/2026.06.19.26356030

**Authors:** Jingjing Gao, Jason H. Windett, Lilian O. Ademu, Zhi Li, Muinat Abolore Idris, Bryan Colby Griffin, Yue Zhang, Benjamin J. Radford

## Abstract

**Background:** Human papillomavirus (HPV) vaccination is an effective cancer prevention strategy, yet HPV vaccine awareness remains uneven across sociodemographic groups. In the current digital information environment, awareness may be shaped not only by access to health information but also by exposure to false or misleading health information, difficulty evaluating information accuracy, and echo-chamber dynamics on social media.

**Objective:** This study examined associations between perceived exposure to false or misleading health information on social media, difficulty determining whether social media health information is true or false, perceived echo-chamber exposure, and HPV vaccine awareness among U.S. adults.

**Methods:** We analyzed nationally representative Health Information National Trends Survey data using survey-weighted descriptive statistics and logistic regression models. The analytic sample included 2,371 respondents, representing a weighted population of 49.2 million U.S. adults. The outcome was HPV vaccine awareness. Primary predictors included perceived exposure to false or misleading health information on social media, difficulty determining whether social media health information was true or false, and perceived same-view health network exposure on social media. Models adjusted for age, sex, race/ethnicity, education, household income, rurality, and Census division.

**Results:** Overall, 60.37% of respondents reported HPV vaccine awareness. Most respondents reported encountering false or misleading health information on social media, with 45.57% reporting “some” and 32.83% reporting “a lot.” In unadjusted models, greater perceived exposure to false or misleading health information was associated with higher odds of HPV vaccine awareness. After adjustment, respondents reporting “some” false or misleading health information had significantly higher odds of HPV vaccine awareness compared with those reporting none (AOR=2.40, 95% CI: 1.06-5.43), while the association for “a lot” was marginal (AOR=2.29, 95% CI: 0.97-5.38). Difficulty identifying true versus false social media health information and perceived echo-chamber exposure were associated with HPV vaccine awareness in unadjusted models but were attenuated after adjustment. HPV vaccine awareness was substantially higher among females and respondents with higher educational attainment, and lower among Hispanic, non-Hispanic Asian, and non-Hispanic other respondents compared with non-Hispanic White respondents.

**Conclusions:** HPV vaccine awareness is associated with both digital health information exposure and persistent sociodemographic inequities. Greater perceived exposure to misleading health information may reflect broader engagement with health-related content on social media, where accurate and inaccurate information coexist. Public health communication strategies should address misinformation vulnerability while expanding accurate, culturally responsive HPV vaccine messaging across digital platforms.

## Introduction

Human papillomavirus (HPV) vaccination is a major cancer prevention strategy, yet public awareness of the HPV vaccine remains uneven across population groups. In the US, awareness of HPV and the HPV vaccine declined between 2008 and 2018, with the lowest awareness among racial minorities, rural residents, men, older adults, and those with lower education and income [1, 2]. Although HPV vaccine communication has traditionally been framed through clinical encounters, school-based vaccination policies, and cervical cancer prevention campaigns, exposure to health information increasingly occurs in digital and social media environments.

These environments can expand access to vaccine-related information, but they also expose users to false, misleading, incomplete, or decontextualized health claims. As a result, HPV vaccine awareness may be shaped not only by access to health information but also by the broader quality, credibility, and social organization of health information encountered online.

Health misinformation has become a central concern in public health communication. Chou, Oh, and Klein (2018) define health misinformation as health-related factual claims that are false because they lack scientific support [3], while Swire-Thompson and Lazer (2020) define misinformation as information that contradicts the scientific consensus [4]. Social media platforms have accelerated the circulation of health misinformation by allowing inaccurate claims, rumors, and conspiracy narratives to spread rapidly through user networks. Prior work has shown that health misinformation can influence public health behaviors, including vaccine attitudes and uptake [3, 5]. This concern became especially visible during the COVID-19 pandemic, when researchers documented widespread misinformation across digital platforms, including Twitter, YouTube, and web-based search results [6–8]. Although COVID-19 intensified public attention to health misinformation, similar dynamics are relevant to other vaccine-related topics, including HPV vaccination.

Fact-checking and corrective information have been proposed as important tools for responding to health misinformation. Historically, fact-checking research focused heavily on political communication, elections, and public statements by political actors [9]. However, the public health domain presents distinct challenges. Health information often involves scientific uncertainty, changing evidence, technical language, and high levels of public anxiety. During health crises, fact-checking must compete with information overload, low trust, and competing interpretations of risk [10, 11]. Even when fact-checks are accurate, their effects may vary across audiences and may not persist over time [9]. Therefore, examining how people perceive the difficulty of determining whether social media health information is true or false is important for understanding misinformation vulnerability in vaccine communication.

Echo chambers further complicate the public health information environment. Social media users may cluster into networks where they encounter information that reinforces existing beliefs, values, and health-related views. These same-view networks can reduce exposure to corrective information and increase selective exposure to content that aligns with prior attitudes [12, 13].

Confirmation bias can also shape how people search for, interpret, and share vaccine-related information, especially when health topics become politically or culturally contested [14]. In vaccine communication, echo chambers may contribute to misinformation persistence by reinforcing doubts about vaccine safety, mistrust of health institutions, or selective acceptance of evidence.

At the same time, exposure to misleading health information does not necessarily indicate lower health awareness. Individuals who are more active in online health information environments may encounter both accurate and inaccurate content. Thus, perceived exposure to misinformation may reflect broader engagement with health-related content on social media rather than misinformation susceptibility alone. This distinction is important for HPV vaccine awareness because social media may simultaneously function as a source of awareness, confusion, correction, and misinformation. Understanding these competing dynamics can help public health practitioners design communication strategies that do not simply increase message volume but also improve information quality, trust, and interpretability.

Prior research also indicates that HPV vaccine awareness is socially patterned across demographic, socioeconomic, and geographic characteristics. Awareness has generally been lower among men, older adults, racial and ethnic minority populations, individuals with lower educational attainment, and adults with lower household income [2, 15–21]. These disparities may reflect differences in provider recommendation, health care access, health literacy, exposure to vaccine-related messaging, language access, and trust in health institutions. Geographic context may also shape HPV vaccine awareness through variation in rurality, preventive care access, school and state vaccine policies, local public health infrastructure, and regional communication environments [22–30]. Because these factors may influence both exposure to social media health information and HPV vaccine awareness, adjustment for sociodemographic and geographic characteristics is important for estimating whether digital misinformation-related factors are associated with HPV vaccine awareness beyond established population differences.

Despite growing concern about digital health misinformation, limited national evidence has examined how perceived exposure to false or misleading health information, difficulty assessing information accuracy, and echo-chamber exposure are associated with HPV vaccine awareness among U.S. adults. This study addresses this gap using nationally representative Health Information National Trends Survey data. We examine whether perceived exposure to false or misleading health information on social media, difficulty determining whether social media health information is true or false, and perceived same-view health network exposure are associated with HPV vaccine awareness. We also assess whether these associations persist after adjustment for sociodemographic and geographic characteristics. By linking HPV vaccine awareness to the digital misinformation environment, this study contributes to public health research on vaccine communication, health equity, and misinformation vulnerability in social media settings.

## Methods

### Study Design and Data Source

This study used a cross-sectional design and analyzed data from the Health Information National Trends Survey (HINTS), a nationally representative survey of U.S. adults that assesses health communication, health information seeking, digital health engagement, cancer-related knowledge, and health-related attitudes. HINTS uses a complex survey design with sampling weights, stratification, and clustering to generate population-representative estimates. The present analysis focused on respondents with valid data on HPV vaccine awareness, social media misinformation exposure, fact-checking difficulty, echo-chamber exposure, and selected sociodemographic and geographic covariates.

### Analytic Sample

The analytic sample was restricted to respondents with complete data for the outcome, primary predictors, and covariates used in the fully adjusted regression model. After applying these criteria, the final analytic sample included 2,371 respondents, representing a weighted population of 49,228,728 U.S. adults. Respondents excluded from the adjusted model were those with missing data on one or more variables included in the primary model.

### Outcome Variable

The primary outcome was HPV vaccine awareness. HPV vaccine awareness was assessed using the HINTS item asking whether respondents had ever heard of the cervical cancer vaccine or HPV shot. Responses were recoded as a binary variable, with “Yes” coded as 1 and “No” coded as 0. Respondents with missing or inapplicable responses were excluded from the analytic model.

### Primary Independent Variables

The primary independent variables captured three dimensions of the social media health information environment: **perceived exposure to false or misleading health information**, **difficulty determining whether social media health information is true or false, and perceived echo-chamber exposure**. *Perceived exposure to false or misleading health information on social media* was measured using a HINTS item asking respondents how much false or misleading health information they saw on social media. Responses were categorized as none, a little, some, and a lot, with “none” used as the reference category in regression models. *Fact-checking difficulty* was measured using an item assessing agreement with the statement that it is difficult to determine whether health information on social media is true or false.

Responses were categorized as strongly disagree, somewhat disagree, somewhat agree, and strongly agree. “Strongly disagree” was used as the reference category. ***Perceived echo-chamber exposure*** was measured using an item assessing whether most people in the respondent’s social media network share the same health views. Responses were categorized as strongly disagree, somewhat disagree, somewhat agree, and strongly agree. “Strongly disagree” was used as the reference category. This measure was used to capture perceived same-view health-information networks on social media.

### Covariates

Multivariable models adjusted for sociodemographic and geographic characteristics selected a priori based on their relevance to HPV vaccine awareness, digital health information exposure, and health communication inequities. Covariates included age, sex, race/ethnicity, educational attainment, household income, rurality, and U.S. Census division. Age was modeled as a continuous variable in years. Sex was modeled as a binary variable, with male as the reference category and female as the comparison group. Race/ethnicity was categorized as non-Hispanic White, non-Hispanic Black, Hispanic, non-Hispanic Asian, and non-Hispanic other, with non-Hispanic White used as the reference category. Educational attainment was categorized as less than high school, high school graduate, some college, bachelor’s degree, and post-baccalaureate degree, with less than high school used as the reference category. Household income was categorized as less than $20,000, $20,000-$34,999, $35,000-$49,999, and $50,000-$74,999, with less than $20,000 used as the reference category. Rurality was derived from the 2013 Rural-Urban Continuum Codes and categorized as metropolitan versus nonmetropolitan/rural, with metropolitan residence used as the reference category. Geographic region was measured using U.S. Census division and categorized as New England, Middle Atlantic, East North Central, West North Central, South Atlantic, East South Central, West South Central, Mountain, and Pacific. South Atlantic was specified as the reference category in regression models.

### Statistical Analysis

All analyses accounted for the complex HINTS survey design using survey weights, strata, and primary sampling units. Weighted descriptive statistics were calculated for all variables in the analytic sample. Categorical variables were summarized using weighted percentages, and age was summarized using the weighted mean. Weighted cross-tabulations were used to describe respondent characteristics by HPV vaccine awareness status, and design-based Pearson chi-square tests were used to assess bivariate differences across categorical variables. Survey-weighted logistic regression models were used to estimate associations between the social media health information environment and HPV vaccine awareness. Model 1 included perceived false or misleading health information exposure, fact-checking difficulty, and perceived echo-chamber exposure. Model 2 additionally adjusted for age, sex, race/ethnicity, education, household income, rurality, and Census division. Results are presented as odds ratios with 95% confidence intervals. An exploratory interaction model was also estimated to assess whether the association between age and HPV vaccine awareness differed by perceived echo-chamber exposure. This model included an interaction term between continuous age and echo-chamber exposure, while retaining the same covariates as the fully adjusted model. Predicted probabilities were generated from the interaction model to visualize age-related differences in HPV vaccine awareness across levels of perceived echo-chamber exposure. Because the interaction analysis was exploratory, it was interpreted cautiously and used to assess potential heterogeneity rather than to define the main study findings. All statistical tests were two-sided, and statistical significance was evaluated at p<0.05. Analyses were conducted using Stata with survey commands to account for the complex sampling design.

### Ethical Considerations

This study used publicly available, de-identified survey data. Because the analysis involved secondary analysis of de-identified public-use data, it did not constitute human subjects research requiring institutional review board approval.

## Results

### Social Media Information Environment and HPV Vaccine Awareness

In the analytic sample, 60.37% of U.S. adults reported HPV vaccine awareness. Most respondents reported encountering at least some false or misleading health information on social media, with 45.57% reporting “some” and 32.83% reporting “a lot.” HPV vaccine awareness was higher among respondents reporting greater perceived exposure to false or misleading health information on social media, increasing from 40.16% among those reporting no exposure to 62.21% among those reporting some exposure and 63.68% among those reporting a lot of exposure; this association was marginally significant in survey-weighted bivariate analysis (p=0.053). Difficulty determining whether social media health information was true or false differed significantly by HPV vaccine awareness status (p=0.018). Awareness was highest among respondents who somewhat disagreed that it was difficult to determine whether social media health information was true or false (70.50%) and lowest among those who somewhat agreed (54.70%). Perceived same-view health network exposure on social media was not significantly associated with HPV vaccine awareness in bivariate analysis (p=0.099). Substantial sociodemographic differences were observed. HPV vaccine awareness was higher among females than males (71.54% vs 46.56%; p<0.001). Awareness also varied significantly by race and ethnicity (p<0.001), with the highest awareness among non-Hispanic White respondents (69.43%) and the lowest among non-Hispanic Asian respondents (26.74%). Higher educational attainment was strongly associated with HPV vaccine awareness (p<0.001), increasing from 43.02% among respondents with less than a high school education to 79.84% among those with post-baccalaureate education. Household income was also significantly associated with awareness (p=0.003). Geographically, HPV vaccine awareness was higher among metropolitan respondents than nonmetropolitan/rural respondents in bivariate analysis (67.90% vs 59.07%; p=0.027). Figure 1 also displays weighted HPV vaccine awareness by U.S. Census division.

**Figure 1.**
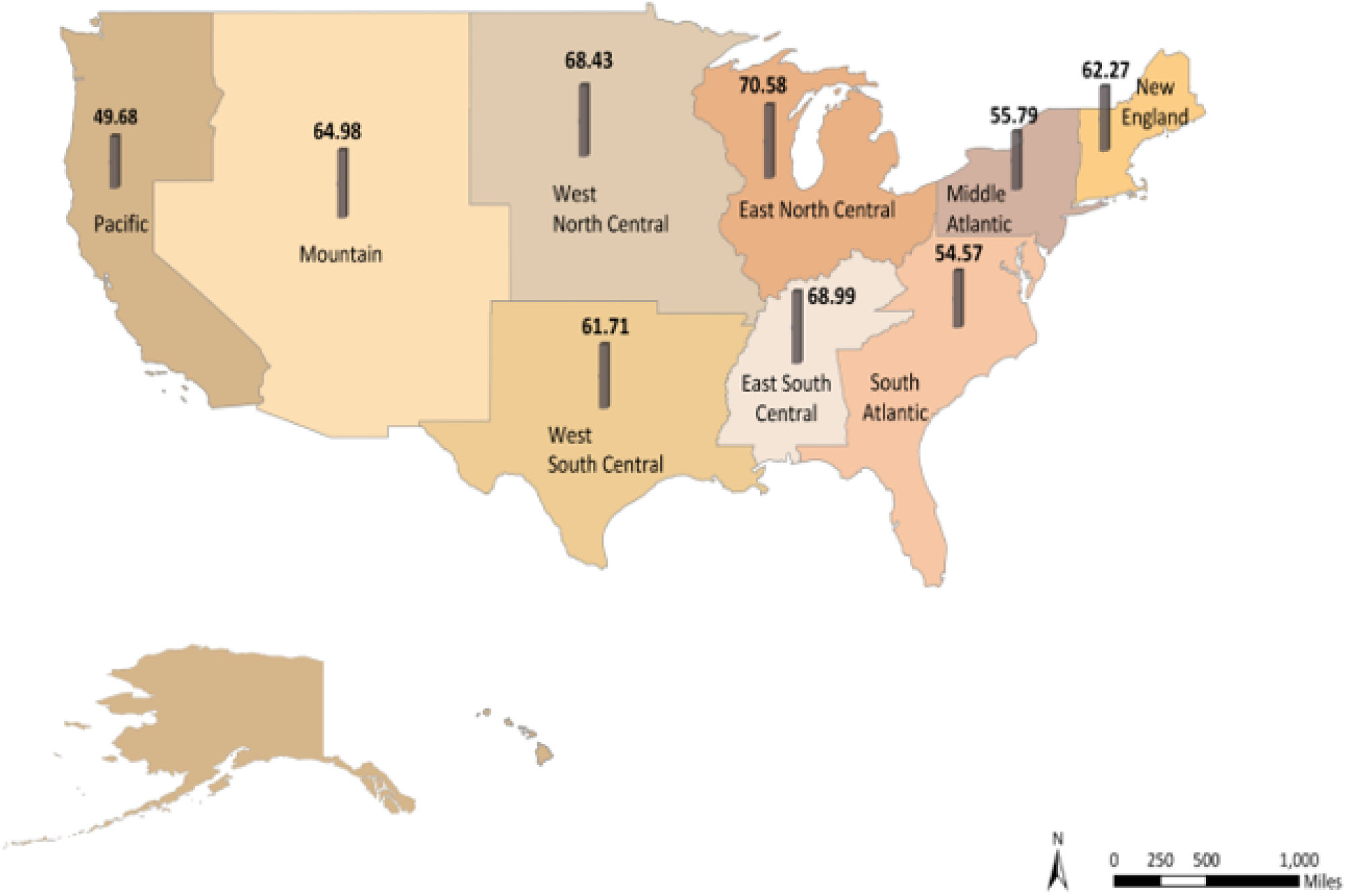
Geographic Variation in HPV Vaccine Awareness by U.S. Census Division. Note. Values represent survey-weighted percentages of adults reporting HPV vaccine awareness within each U.S. Census division. HPV vaccine awareness was defined as having heard of the cervical cancer vaccine or HPV shot.

Descriptively, awareness was highest in the East North Central division (70.58%), followed by the East South Central (68.99%), West North Central (68.43%), and Mountain (64.98%) divisions. Awareness was lowest in the Pacific division (49.68%), South Atlantic division (54.57%), and Middle Atlantic division (55.79%). Although the map shows visible regional variation in HPV vaccine awareness, the overall difference by Census division was not statistically significant in survey-weighted bivariate analysis (p=0.101). These findings suggest that regional patterns may be present descriptively but should be interpreted cautiously.

**Table 1.**
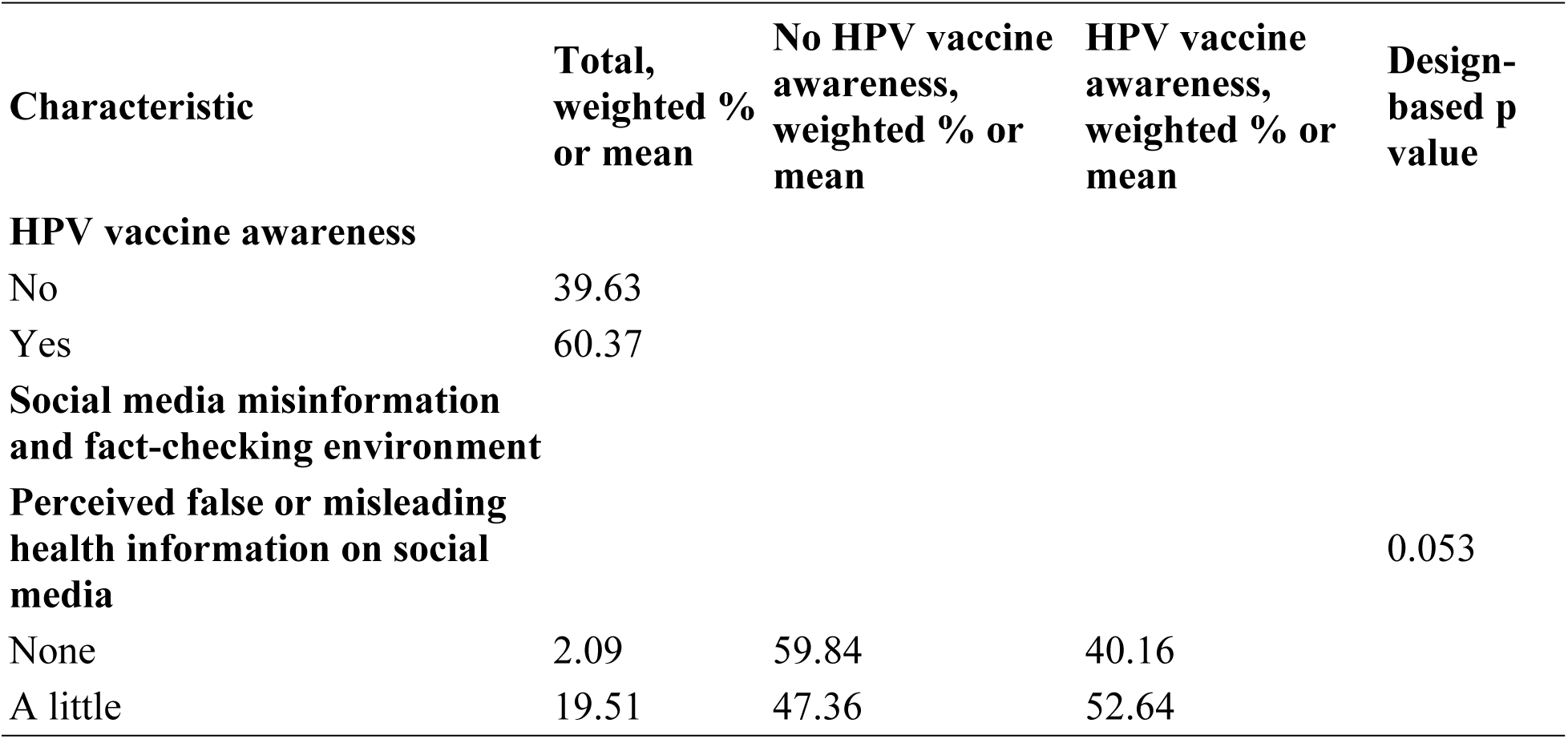
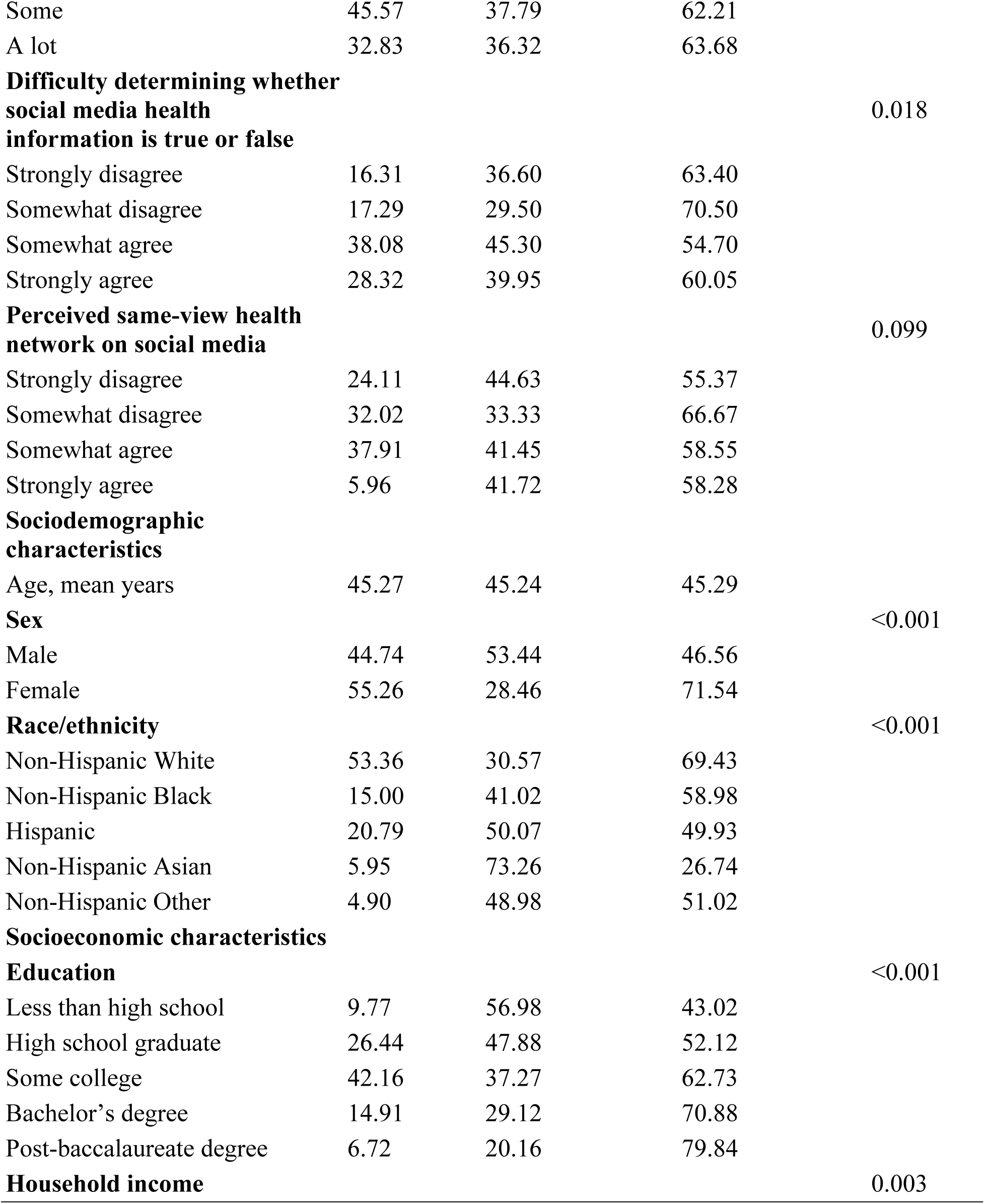
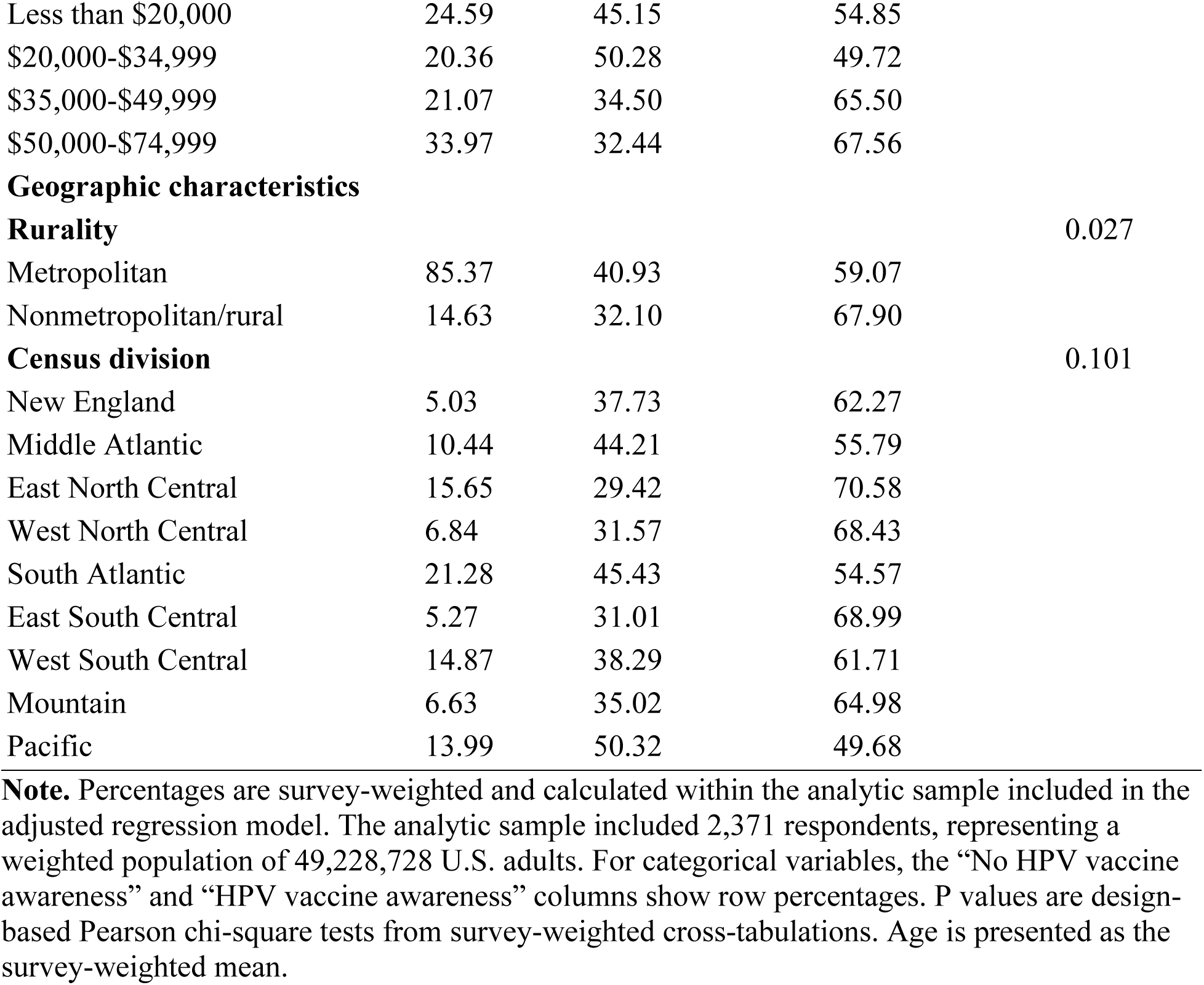
Weighted descriptive characteristics of U.S. adults included in the HPV vaccine awareness and social media misinformation analysis

### Associations Between Social Media Misinformation Exposure, Fact-Checking Difficulty, Echo-Chamber Exposure, and HPV Vaccine Awareness

In survey-weighted logistic regression models, perceived exposure to false or misleading health information on social media was associated with HPV vaccine awareness. In the unadjusted model, compared with respondents who reported seeing no false or misleading health information on social media, those reporting “a little” misleading information had higher odds of HPV vaccine awareness (OR=2.19, 95% CI: 1.16-4.14), as did those reporting “some” (OR=3.27, 95% CI: 1.76-6.09) and “a lot” (OR=4.45, 95% CI: 2.32-8.53). After adjustment for sociodemographic and geographic characteristics, the association remained statistically significant for respondents reporting “some” misleading health information (AOR=2.40, 95% CI: 1.06-5.43), while the association for “a lot” was marginal (AOR=2.29, 95% CI: 0.97-5.38).

Difficulty determining whether social media health information was true or false was inversely associated with HPV vaccine awareness in the unadjusted model. Respondents who somewhat agreed (OR=0.59, 95% CI: 0.41-0.86) or strongly agreed (OR=0.72, 95% CI: 0.54-0.96) that it was difficult to determine whether social media health information was true or false had lower odds of HPV vaccine awareness than those who strongly disagreed. These associations were attenuated and no longer statistically significant after adjustment.

Perceived echo-chamber exposure showed a positive association with HPV vaccine awareness in the unadjusted model among respondents who somewhat disagreed (OR=1.58, 95% CI: 1.15-2.17) or somewhat agreed (OR=1.38, 95% CI: 1.01-1.89) that most people in their social media networks shared their health views. However, these associations were not statistically significant in the adjusted model. In the exploratory interaction model, the main effect for somewhat disagreeing with same-view health networks was positive (OR=3.66, 95% CI: 1.02-13.12), while the age interaction for this category was marginally negative (OR=0.98, 95% CI: 0.95-1.00), suggesting possible age-related heterogeneity in the association between echo-chamber exposure and HPV vaccine awareness.

Several sociodemographic differences were observed. Female respondents had more than three times the odds of HPV vaccine awareness compared with male respondents in the adjusted model (AOR=3.42, 95% CI: 2.48-4.73). Compared with non-Hispanic White respondents, Hispanic respondents (AOR=0.42, 95% CI: 0.26-0.66), non-Hispanic Asian respondents (AOR=0.12, 95% CI: 0.06-0.27), and non-Hispanic other respondents (AOR=0.39, 95% CI: 0.19-0.80) had significantly lower odds of HPV vaccine awareness. Higher educational attainment was positively associated with awareness, particularly among respondents with a bachelor’s degree (AOR=2.31, 95% CI: 1.26-4.25) or post-baccalaureate degree (AOR=3.51, 95% CI: 1.79-6.88). Rurality was not significantly associated with HPV vaccine awareness after adjustment.

**Table 2.** Survey-weighted logistic regression models of HPV vaccine awareness by social media misinformation exposure, fact-checking difficulty, and echo-chamber exposure among U.S. adults

| Variable | Model 1:<br>Unadjusted OR<br>(95% CI) | Model 2:<br>Adjusted OR<br>(95% CI) | Model 3:<br>Age × echo chamber<br>interaction OR (95%<br>CI) |
| --- | --- | --- | --- |
| **Perceived false/misleading health information on social media** |  |  |  |
| Reference: None |  |  |  |
| A little | 2.19 (1.16-4.14)* | 1.47 (0.63-3.42) | 1.42 (0.61-3.33) |
| Some | 3.27 (1.76-6.09)*** | 2.40 (1.06-5.43)* | 2.43 (1.08-5.48)* |
| A lot | 4.45 (2.32-8.53)*** | 2.29 (0.97-5.38) | 2.26 (0.96-5.29) |
| **Difficulty determining whether social media health information is true or false** |  |  |  |
| Reference: Strongly disagree |  |  |  |
| Somewhat disagree | 0.86 (0.55-1.34) | 1.38 (0.81-2.34) | 1.38 (0.81-2.34) |
| Somewhat agree | 0.59 (0.41-0.86)** | 0.65 (0.39-1.09) | 0.65 (0.39-1.08) |
| Strongly agree | 0.72 (0.54-0.96)* | 0.67 (0.40-1.11) | 0.66 (0.40-1.10) |
| **Perceived health-information**<br>**echo chamber exposure** |  |  |  |
| Reference: Strongly disagree |  |  |  |
| Somewhat disagree | 1.58 (1.15-2.17)** | 1.22 (0.82-1.84) | 3.66 (1.02-13.12)* |
| Somewhat agree | 1.38 (1.01-1.89)* | 1.04 (0.65-1.69) | 0.91 (0.24-3.49) |
| Strongly agree | 1.17 (0.72-1.90) | 0.87 (0.47-1.62) | 0.77 (0.14-4.26) |
| **Age, years** |  | 0.99 (0.98-1.00)* | 0.99 (0.98-1.01) |
| **Age × echo chamber exposure** |  |  |  |
| Age × Somewhat disagree |  |  | 0.98 (0.95-1.00) |
| Age × Somewhat agree |  |  | 1.00 (0.98-1.03) |
| Age × Strongly agree |  |  | 1.00 (0.97-1.03) |
| **Female** |  | 3.42 (2.48-4.73)*** | 3.39 (2.44-4.72)*** |
| **Race/ethnicity** |  |  |  |
| Reference: Non-Hispanic White |  |  |  |
| Non-Hispanic Black |  | 0.62 (0.37-1.02) | 0.61 (0.37-1.03) |
| Hispanic |  | 0.42 (0.26-0.66)*** | 0.41 (0.25-0.65)*** |
| Non-Hispanic Asian |  | 0.12 (0.06-0.27)*** | 0.12 (0.06-0.26)*** |
| Non-Hispanic Other |  | 0.39 (0.19-0.80)* | 0.38 (0.19-0.76)** |
| **Education** |  |  |  |
| Reference: Less than high school |  |  |  |
| High school graduate |  | 1.06 (0.59-1.90) | 1.06 (0.60-1.86) |
| Some college |  | 1.72 (0.96-3.06) | 1.73 (0.98-3.05) |
| Bachelor's degree |  | 2.31 (1.26-4.25)** | 2.34 (1.28-4.30)** |
| Post-baccalaureate degree |  | 3.51 (1.79-6.88)*** | 3.48 (1.79-6.77)*** |
| **Household income** |  |  |  |
| Reference: Less than \$20,000 | | | |
| \$20,000-\$34,999 | | 0.77 (0.48-1.23) | 0.76 (0.48-1.23) |
| \$35,000-\$49,999 | | 1.62 (1.03-2.56)* | 1.56 (0.99-2.45) |

| **Variable** | **Model 1:**<br>**Unadjusted OR**<br>**(95% CI)** | **Model 2:**<br>**Adjusted OR**<br>**(95% CI)** | **Model 3:**<br>**Age × echo chamber**<br>**interaction OR (95%**<br>**CI)** |
| --- | --- | --- | --- |
| \$50,000-\$74,999 | | 1.70 (1.08-2.69)* | 1.72 (1.08-2.72)* |
| **Rurality** |  |  |  |
| Reference: Metropolitan |  |  |  |
| Nonmetropolitan/rural |  | 1.24 (0.82-1.89) | 1.23 (0.81-1.87) |
| **Census division** |  |  |  |
| Reference: South Atlantic |  |  |  |
| New England |  | 1.05 (0.45-2.47) | 1.10 (0.46-2.64) |
| Middle Atlantic |  | 1.30 (0.71-2.38) | 1.29 (0.72-2.32) |
| East North Central |  | 2.24 (1.30-3.84)** | 2.27 (1.32-3.90)** |
| West North Central |  | 2.08 (0.88-4.87) | 2.12 (0.92-4.86) |
| East South Central |  | 2.18 (1.14-4.19)* | 2.17 (1.13-4.18)* |
| West South Central |  | 1.79 (1.01-3.17)* | 1.81 (1.02-3.24)* |
| Mountain |  | 2.11 (1.07-4.16)* | 2.11 (1.07-4.19)* |
| Pacific |  | 1.35 (0.81-2.26) | 1.39 (0.84-2.30) |
| Observations | 4,510 | 2,371 | 2,371 |
| Weighted population | 100,072,995 | 49,228,728 | 49,228,728 |
| Model F test | F(9,188)=5.46,<br>p<.001 | F(31,161)=7.38,<br>p<.001 | F(34,158)=6.78,<br>p<.001 |

Note: OR = odds ratio; CI = confidence interval. Model 1 included social media misinformation exposure, fact-checking difficulty, and echo-chamber exposure. Model 2 additionally adjusted for age, sex, race/ethnicity, education, household income, rurality, and Census division, with South Atlantic as the reference region. Model 3 added the age × echo-chamber exposure interaction. Reference categories were no perceived false/misleading health information, strongly disagree for fact-checking difficulty, strongly disagree for echo-chamber exposure, male sex, non-Hispanic White race/ethnicity, less than high school education, household income <\$20,000, metropolitan residence, and the South Atlantic Census division. \*p<0.05; \*\*p<0.01; \*\*\*p<0.001.

### Exploratory Age-by-Echo-Chamber Interaction

Figure 2 presents the predicted probability of HPV vaccine awareness by age across levels of perceived health-information echo-chamber exposure. Overall, the predicted probability of HPV vaccine awareness declined modestly with age among respondents who strongly disagreed that most people in their social media networks shared their health views. A steeper age-related decline was observed among respondents who somewhat disagreed with same-view health network exposure, with predicted awareness decreasing from approximately 0.77 among younger adults to approximately 0.44 among older adults. In contrast, predicted probabilities were relatively stable across age among respondents who somewhat agreed or strongly agreed that their social media networks reflected similar health views.

**Figure 2.**
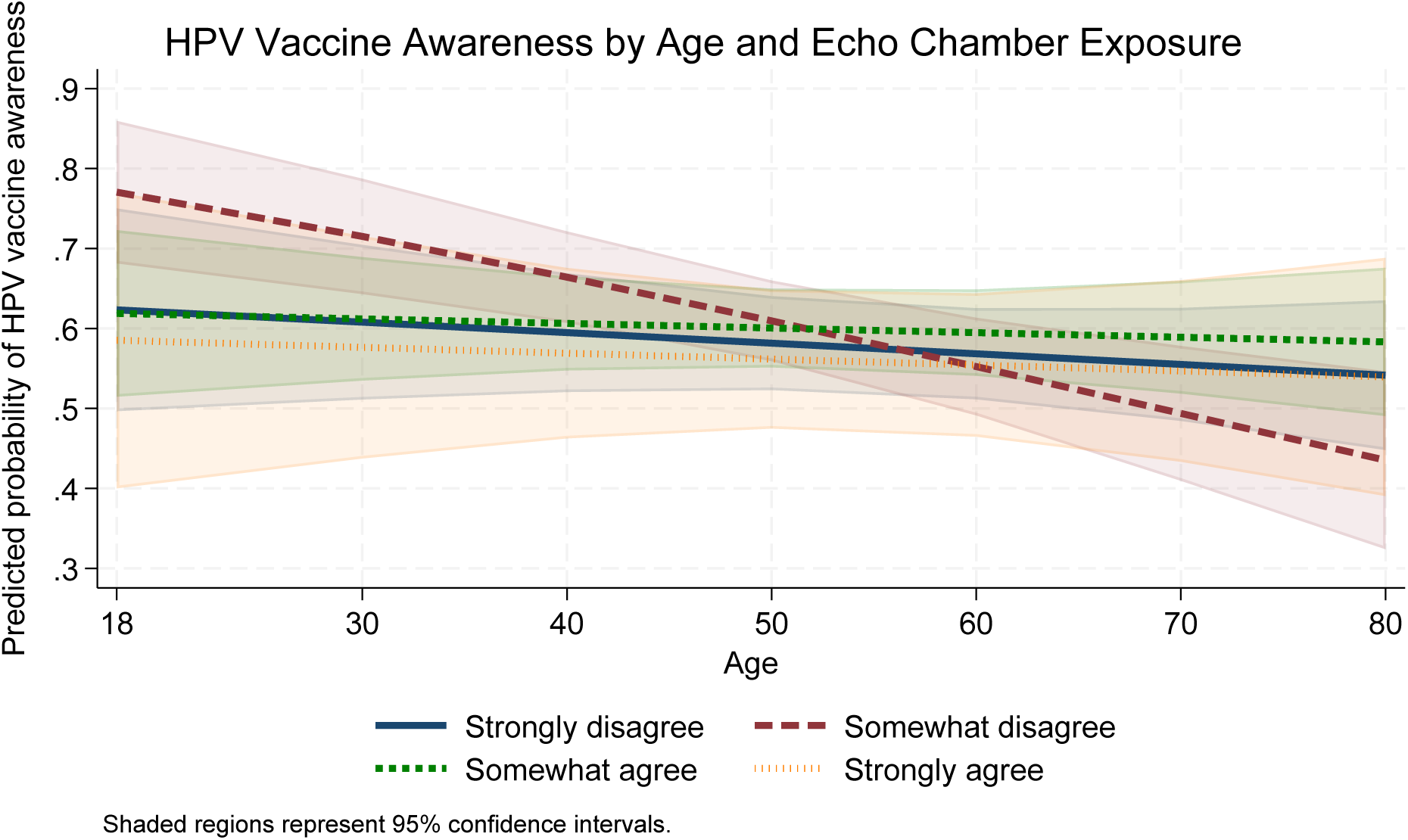
Predicted Probability of HPV Vaccine Awareness by Age and Perceived Health-Information Echo-Chamber Exposure

The shaded 95% confidence intervals indicate substantial uncertainty and overlap across echo-chamber categories, particularly at younger and older ages. Consistent with the regression model, the age interaction for the “somewhat disagree” echo-chamber category was marginally negative, suggesting possible age-related heterogeneity in the association between echo-chamber exposure and HPV vaccine awareness. However, because confidence intervals overlapped and the interaction pattern was exploratory, these findings should be interpreted cautiously. The figure suggests that age-related differences in HPV vaccine awareness may vary across perceived social media health-information environments, but the main adjusted findings remain centered on misinformation exposure and sociodemographic disparities.

### Discussion Principal Findings

This study examined the association between the social media health information environment and HPV vaccine awareness among U.S. adults. Three main findings emerged. First, perceived exposure to false or misleading health information on social media was associated with HPV vaccine awareness. In adjusted models, respondents who reported seeing “some” false or misleading health information on social media had significantly higher odds of HPV vaccine awareness compared with respondents who reported seeing none, while the association among those reporting “a lot” of false or misleading information was positive but marginal. Second, difficulty determining whether social media health information was true or false and perceived echo-chamber exposure were associated with HPV vaccine awareness in unadjusted models, but these associations were attenuated after adjustment for sociodemographic and geographic characteristics. Third, HPV vaccine awareness varied substantially by sex, race/ethnicity, and educational attainment, indicating persistent inequities in vaccine-related awareness.

The positive association between perceived exposure to false or misleading health information and HPV vaccine awareness should be interpreted cautiously. This finding does not suggest that misinformation improves health awareness or should be considered beneficial. Rather, it likely reflects broader engagement with health-related content on social media. Individuals who are more exposed to health information online may be more likely to encounter both accurate and inaccurate content. In this context, perceived exposure to misinformation may serve as a marker of greater overall exposure to social media health information. For HPV vaccination, this distinction is important because digital platforms may simultaneously increase vaccine awareness, amplify uncertainty, expose users to misleading claims, and create opportunities for correction.

These findings contribute to digital public health research by showing that HPV vaccine awareness is connected to the broader social media information environment. Prior work has emphasized that health misinformation can spread rapidly online and may influence vaccine attitudes, health behaviors, and trust in public health institutions [31–33]. The present study extends this literature by focusing on HPV vaccine awareness rather than misinformation belief or vaccine hesitancy alone. Awareness is an early and necessary step in the pathway toward vaccine knowledge, intention, and uptake. Therefore, understanding how social media misinformation exposure relates to awareness can help public health practitioners identify both risks and opportunities in digital vaccine communication.

### Fact-Checking Difficulty and Misinformation Vulnerability

Respondents who reported greater difficulty determining whether social media health information was true or false had lower odds of HPV vaccine awareness in unadjusted models. However, these associations were no longer statistically significant after adjustment. This attenuation suggests that fact-checking difficulty may be intertwined with sociodemographic factors, educational attainment, and broader differences in health information access [34–38]. Individuals with fewer resources for evaluating online health information may be more vulnerable to misinformation, but this vulnerability may operate through structural and informational inequities rather than through individual-level digital skills alone.

This finding has implications for public health communication. Efforts to address misinformation should not rely only on correcting false claims after they appear. Public health agencies and health systems should also strengthen digital health literacy, improve the visibility of credible sources, and design messages that help users evaluate the credibility of online health content. For HPV vaccination specifically, communication strategies should provide clear, accessible, and culturally relevant information about the vaccine, its cancer prevention benefits, and its relevance to all eligible populations, including both males and females.

### Echo-Chamber Exposure and Age-Related Patterns

Perceived same-view health network exposure showed positive associations with HPV vaccine awareness in unadjusted models, but these associations were not significant after adjustment. This suggests that echo-chamber exposure alone may not independently explain HPV vaccine awareness once demographic and geographic differences are considered. However, exploratory interaction results suggested possible age-related heterogeneity. The predicted probability plot indicated that HPV vaccine awareness declined more steeply with age among respondents who somewhat disagreed that most people in their social media networks shared their health views, while predicted awareness was relatively stable across age among other echo-chamber categories.

These exploratory findings should be interpreted with caution because the confidence intervals overlapped across groups. Still, they suggest that age may shape how individuals experience and interpret social media health information environments. Younger adults may have greater exposure to HPV vaccine messaging through social media, educational settings, or peer networks, whereas older adults may be less likely to encounter HPV vaccine information or may associate HPV vaccination primarily with younger populations, which is consistent with the previous studies [39–41]. Future research should examine whether age modifies the relationship between online health information exposure, misinformation vulnerability, and vaccine awareness using longitudinal or experimental designs.

Sociodemographic and Equity Implications

The biggest and most consistent differences in HPV vaccine awareness were observed across sociodemographic groups. Female respondents had substantially higher odds of HPV vaccine awareness compared with male respondents [16, 42]. This pattern is consistent with the historical framing of HPV vaccination as a cervical cancer prevention strategy. Although this framing has been effective in emphasizing the prevention of cervical cancer, it may have unintentionally contributed to lower awareness among males. HPV vaccination is relevant for preventing multiple HPV-associated cancers, including anal, penile, and oropharyngeal cancers. Public health messaging should therefore frame HPV vaccination as a cancer prevention strategy for all eligible individuals, not only as a women’s health issue.

Racial and ethnic disparities were also evident. Hispanic, non-Hispanic Asian, and non-Hispanic other respondents had significantly lower odds of HPV vaccine awareness than non-Hispanic White respondents after adjustment [2, 16, 43]. These findings point to the need for culturally and linguistically appropriate HPV vaccine communication. Lower awareness among some racial and ethnic groups may reflect differences in access to preventive care, provider recommendations, health information exposure, language access, trust, or the relevance of existing public health campaigns. Digital communication strategies should be designed with attention to these inequities rather than assuming that social media outreach reaches all populations equally.

Educational attainment was positively associated with HPV vaccine awareness. Respondents with bachelor’s and post-baccalaureate education had significantly higher odds of awareness than those with less than a high school education, consistent with previous studies [15, 17, 18, 44–46]. This finding underscores the importance of health literacy and access to understandable vaccine information. HPV vaccine messages should avoid technical language and should be tailored for audiences with varying levels of health literacy. Communication strategies that combine plain language, trusted messengers, visual formats, and community-based dissemination may help reduce awareness gaps.

Public Health Implications

The findings have several implications for public health practice. First, public health agencies should treat social media as both a risk environment and an opportunity for HPV vaccine communication. The same platforms that expose users to misleading health information can also be used to disseminate accurate, engaging, and targeted vaccine messages. Second, misinformation interventions should move beyond fact-checking alone. Although fact-checking remains important, users also need tools and skills to identify credible information, recognize misleading claims, and understand why HPV vaccination matters. Third, HPV vaccine communication should be equity-centered. Messaging should be tailored to groups with lower awareness, including males, racial and ethnic minority populations, and adults with lower educational attainment.

These results also support the importance of monitoring the digital information environment as part of vaccine communication planning. Public health surveillance often focuses on vaccine uptake, hesitancy, or provider recommendation, but awareness is also a critical upstream indicator. Monitoring misinformation exposure, perceived difficulty evaluating online health information, and echo-chamber dynamics may help identify populations at risk of lower awareness or greater confusion about HPV vaccination.

Strengths and Limitations

This study has several strengths. It used nationally representative HINTS data and accounted for the complex survey design, allowing estimates to represent U.S. adults in the analytic sample.

The study also examined multiple dimensions of the social media health information environment, including exposure to misinformation, difficulty with fact-checking, and exposure to echo chambers. In addition, the analysis incorporated sociodemographic and geographic covariates, allowing assessment of whether digital information factors were associated with HPV vaccine awareness beyond key population characteristics.

Several limitations should also be noted. First, the cross-sectional design prevents causal inference. The findings cannot determine whether social media misinformation exposure increases HPV vaccine awareness, whether individuals with greater HPV vaccine awareness are more likely to notice misinformation, or whether both reflect broader engagement with health information. Second, all measures were self-reported and may be subject to recall bias or perception bias. Respondents’ perceived exposure to false or misleading health information may not correspond to objectively verified misinformation exposure. Third, the outcome measured HPV vaccine awareness, not HPV vaccine uptake, intention, or accurate knowledge. Awareness is important, but it does not necessarily translate into vaccination behavior. Fourth, the analytic sample was restricted to respondents with complete data in the adjusted model, which may limit generalizability if missingness was not random. Finally, echo-chamber exposure was measured using perceived same-view health networks, which may not fully capture algorithmic curation, network structure, or actual exposure to homogeneous content.

## Future Research

Future studies should examine whether social media misinformation exposure is associated with HPV vaccine knowledge, vaccine confidence, vaccination intentions, and actual vaccine uptake. Longitudinal studies are needed to clarify directionality and determine whether digital health information exposure predicts later changes in vaccine awareness or behavior. Experimental studies could assess whether corrective information, digital literacy prompts, or culturally tailored HPV vaccine messages improve awareness and reduce misinformation vulnerability. Future research should also examine platform-specific dynamics, including differences across social media platforms, video-sharing sites, search engines, and messaging apps.

## Conclusion

HPV vaccine awareness among U.S. adults is associated with the social media health information environment and marked by substantial sociodemographic inequities. Respondents who reported seeing some false or misleading health information on social media had higher adjusted odds of HPV vaccine awareness, likely reflecting broader exposure to health-related content online rather than a beneficial effect of misinformation. Fact-checking difficulty and perceived echo-chamber exposure were associated with awareness in unadjusted models but were attenuated after adjustment. Awareness was substantially higher among females and adults with higher educational attainment, and lower among Hispanic, non-Hispanic Asian, and non-Hispanic other respondents compared with non-Hispanic White respondents. Public health strategies should address misinformation vulnerability while expanding accurate, culturally responsive, and equity-centered HPV vaccine communication across digital platforms.

## Data Availability

No new data were generated for this study. This study used publicly available, de-identified data from the National Cancer Institute Health Information National Trends Survey (HINTS). The HINTS public-use datasets are available from the National Cancer Institute at https://hints.cancer.gov/data/download-data.aspx. The present analysis used HINTS data with variables on HPV vaccine awareness, social media health information exposure, fact-checking difficulty, perceived same-view health networks, and sociodemographic and geographic characteristics. Analytic code may be made available by the corresponding author upon reasonable request.

## Acknowledgments

Not Available.

## Data Availability

The data analyzed in this study are publicly available from the National Cancer Institute Health Information National Trends Survey website: https://hints.cancer.gov/data/download-data.aspx. The present study used de-identified public-use HINTS data. Analytic code may be available from the corresponding author upon reasonable request.

## Authors’ Contributions

Conceptualization: JG, JHW, BJR Data curation: JG, ZY

Formal analysis: JG, ZY

Investigation: JG, JHW, LOA, ZL, MAI, BCG, ZY, BJR

Methodology: JG, ZY, JHW, BJR Project administration: JG Resources: JG

Software: JG, ZY Supervision: JG Validation: JG, ZY Visualization: JG

Writing - original draft: JG

Writing - review and editing: JG, JHW, LOA, ZL, MAI, BCG, ZY, BJR All authors read and approved the final manuscript.

